# Passive Detection of COVID-19 with Wearable Sensors and Explainable Machine Learning Algorithms

**DOI:** 10.1101/2021.08.05.21261626

**Authors:** Matteo Gadaleta, Jennifer M. Radin, Katie Baca-Motes, Edward Ramos, Vik Kheterpal, Eric J. Topol, Steven R. Steinhubl, Giorgio Quer

**Author notes:** Correspondence to: Giorgio Quer, Scripps Research Translational Institute, 3344 N Torrey Pines Ct Plaza Level, La Jolla CA 92037 USA;.

## Abstract

Individual smartwatch or fitness band sensor data in the setting of COVID-19 has shown promise to identify symptomatic and pre-symptomatic infection or the need for hospitalization, correlations between peripheral temperature and self-reported fever, and an association between changes in heart-rate-variability and infection. In our study, a total of 38,911 individuals (61% female, 15% over 65) have been enrolled between March 25, 2020 and April 3, 2021, with 1,118 reported testing positive and 7,032 negative for COVID-19 by nasopharyngeal PCR swab test. We propose an explainable gradient boosting prediction model based on decision trees for the detection of COVID-19 infection that can adapt to the absence of self-reported symptoms and to the available sensor data, and that can explain the importance of each feature and the post-test-behavior for the individuals. We tested it in a cohort of symptomatic individuals who exhibited an AUC of 0.83 [0.81-0.85], or AUC=0.78 [0.75-0.80] when considering only data before the test date, outperforming state-of-the-art algorithm in these conditions. The analysis of all individuals (including asymptomatic and pre-symptomatic) when self-reported symptoms were excluded provided an AUC of 0.78 [0.76-0.79], or AUC of 0.70 [0.69-0.72] when considering only data before the test date. Extending the use of predictive algorithms for detection of COVID-19 infection based only on passively monitored data from any device, we showed that it is possible to scale up this platform and apply the algorithm in other settings where self-reported symptoms can not be collected.

## INTRODUCTION

Frequent monitoring to quickly identify, trace and isolate cases of SARS-CoV-2 is needed to help control the spread of the infection as well as improve individual patient care through the earlier initiation of effective therapies.^1^ Frequent diagnostic testing is one important option but suffers from implementation challenges and a lack of accessibility for individuals affected most by COVID-19.^2^ Self-reporting of symptoms has been found to be predictive of a positive test,^3^ and could be used to encourage individuals to get tested earlier. However, such an approach not only requires active engagement of the individual, but also misses the approximately one-third of asymptomatic infected individuals completely, and delays diagnosis in those who are infected but presymptomatic.^4^ On the other hand, passive monitoring is possible with commercial sensor devices measuring biometrics such as resting heart rate,^5^ sleep^6^ or activity, which have been shown to be effective in the detection of COVID-19 versus non-COVID-19 when incorporated in combination with self-reported symptoms.^7^

Individual sensor data in the setting of COVID-19 has also shown promise in identifying pre-symptomatic infection,^8^ the need for hospitalization,^9^ correlations between peripheral temperature and self-reported fever,^10^ differences in the changes in wearable data between individuals with COVID-19 versus influenza-like-illnesses,^11^ and an association between changes in heart-rate-variability and infection.^12^ These studies focused on a specific device brand, or on a predefined set of signals. However, for a broader use of personal health technologies it is important to design algorithms that are device agnostic and can adapt to the specific data collected by any sensor, including the less costly devices.

Our prospective app-based research platform DETECT (Digital Engagement and Tracking for Early Control and Treatment) allows participants to enter self-reported symptoms or COVID-19 test results, and to share data from any wearable device that is connected to Google Fit or Apple Health Kit platform. In a previous study, we developed a deterministic algorithm to discriminate between symptomatic individuals testing positive or negative for COVID-19, analyzing changes in daily values of resting heart rate, length of sleep and amount of activity, together with self-reported symptoms.^7^

In order to provide the most accurate early warnings for COVID-19 to all participants for a wide variety of wearable devices, we proposed and validated a machine learning algorithm that ingests all available sensor data for the detection of COVID-19 infection. The algorithm can outperform our previously proposed algorithm in similar conditions (AUC=0.83, IQR=[0.81, 0.85]), and more importantly, it can automatically adapt to the specific sensor used, exploiting all the information collected from the more advanced sensors or focusing on a smaller set of signals from more basic sensors, and explaining the feature importance and the post-test behavioral changes for the individual. The algorithm uses self-reported symptoms when they are available, or otherwise makes its inference based on sensor data only, thus adapting to different engagement levels of the individuals in the study.

## RESULTS

In this study, we investigated the accuracy of a machine learning model in the detection of COVID-19 infection based on the available data acquired from wearable devices and self-reported surveys. We analyzed the accuracy of the detection algorithm for individuals who self-reported at least one symptom prior to the COVID-19 test (named the “symptomatic cohort” in what follows) and not-reporting any symptom prior to the COVID-19 test (the “no-symptom-reported cohort”). We also separately investigated the accuracy obtained using only data collected before a COVID-19 test versus including pre-and post-test data, in order to explore the effect of behavioral changes just the act of testing for COVID-19 might have on individuals.

### Participant characteristics

A total of 38,911 individuals (61% female, 15% over 65) have been enrolled between March 25, 2020 and April 3, 2021. Among these participants, 1,118 (66% female, 8% over 65) reported at least one positive and 7,032 (63% female, 14% over 65) at least one negative COVID-19 nasal swab test. The total number of COVID-19 swab tests reported during the same period was 18,175, with 1,360 (7.5%) positives, 16,398 negatives and 417 with non-reported results. Among the positive tests, 539 (48% of the considered cases) reported at least one symptom in the 15 days preceding the test date, 592 (52%) did not report any symptom, and 229 have been excluded from the analysis for lack of sufficient data or for being too close to a prior test.

### Dataset description

The participants of the study shared their personal device data (including historical data collected prior to enrollment), self-reported symptoms and diagnostic test results during the data collection period. We divided the measures into four categories: symptom features, including all self-reported symptoms; sensor features, including all measures related to activity, heart rate or sleep; anthropometrics; and demographics. (Table 1)

**Table 1 -.**
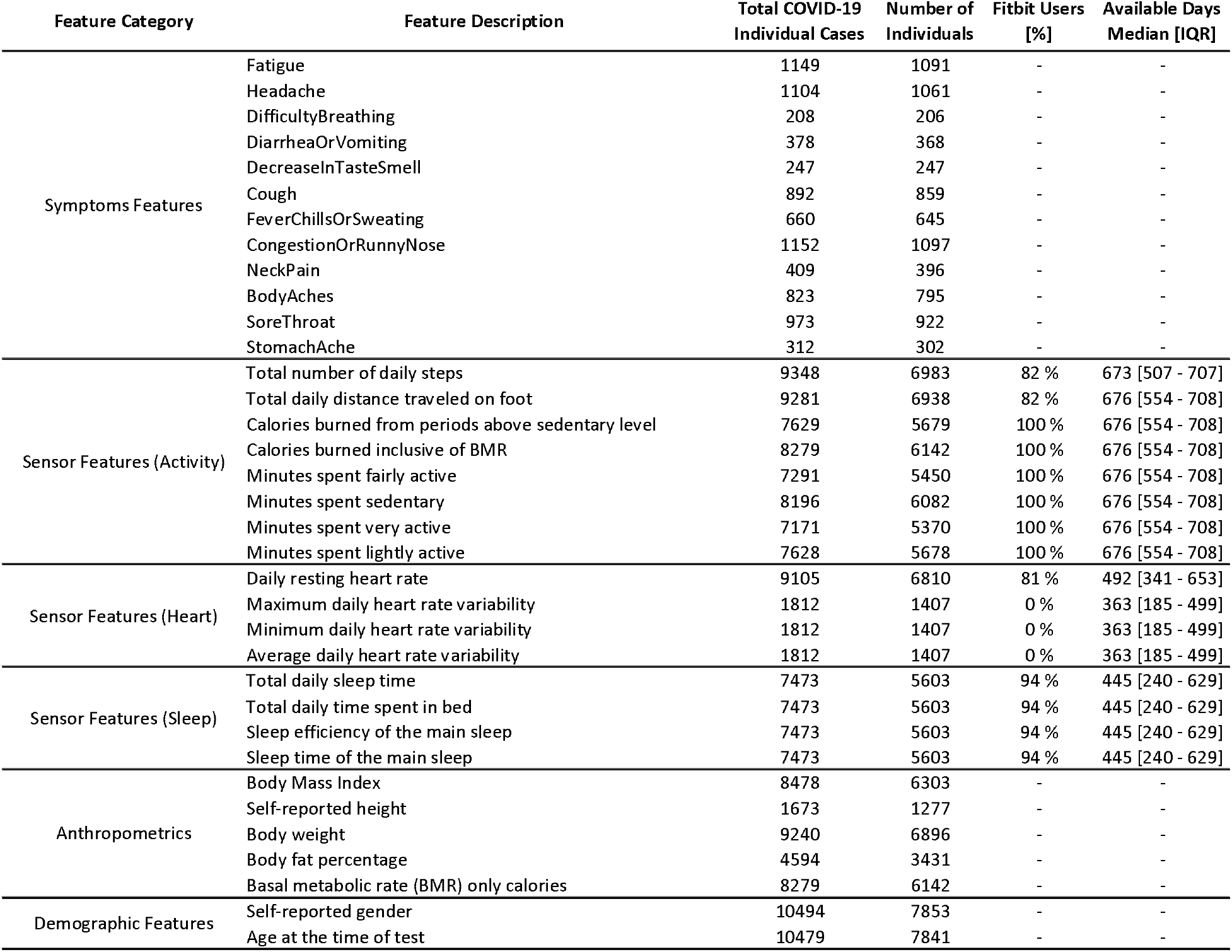
Description and categorization of the features. In the table, one case refers to a specific test from an individual, while an individual may report multiple tests. The total number of individual COVID-19 cases is the number of cases with the corresponding feature value available for the analysis. The available days for Sensor Features represent the median number of days available for all the participants (IQR reported in brackets).

### Detection of COVID-19

The normalized deviations from the baseline for a subset of representative features are reported, (Table 2), highlighting the difference between positive and negative COVID-19 individuals, both excluding and including data after the test date, based on gender and age. As expected, we observed larger variation from the baseline, in terms of heart rate, sleep and activity related features, for individuals who tested positive for COVID-19 with respect to individuals who tested negative. This observation held for all the demographic groups, both excluding or including post-test data. (Figure 1) Based on these features, a prediction model was trained and tested in different conditions.

**Table 2 –.**
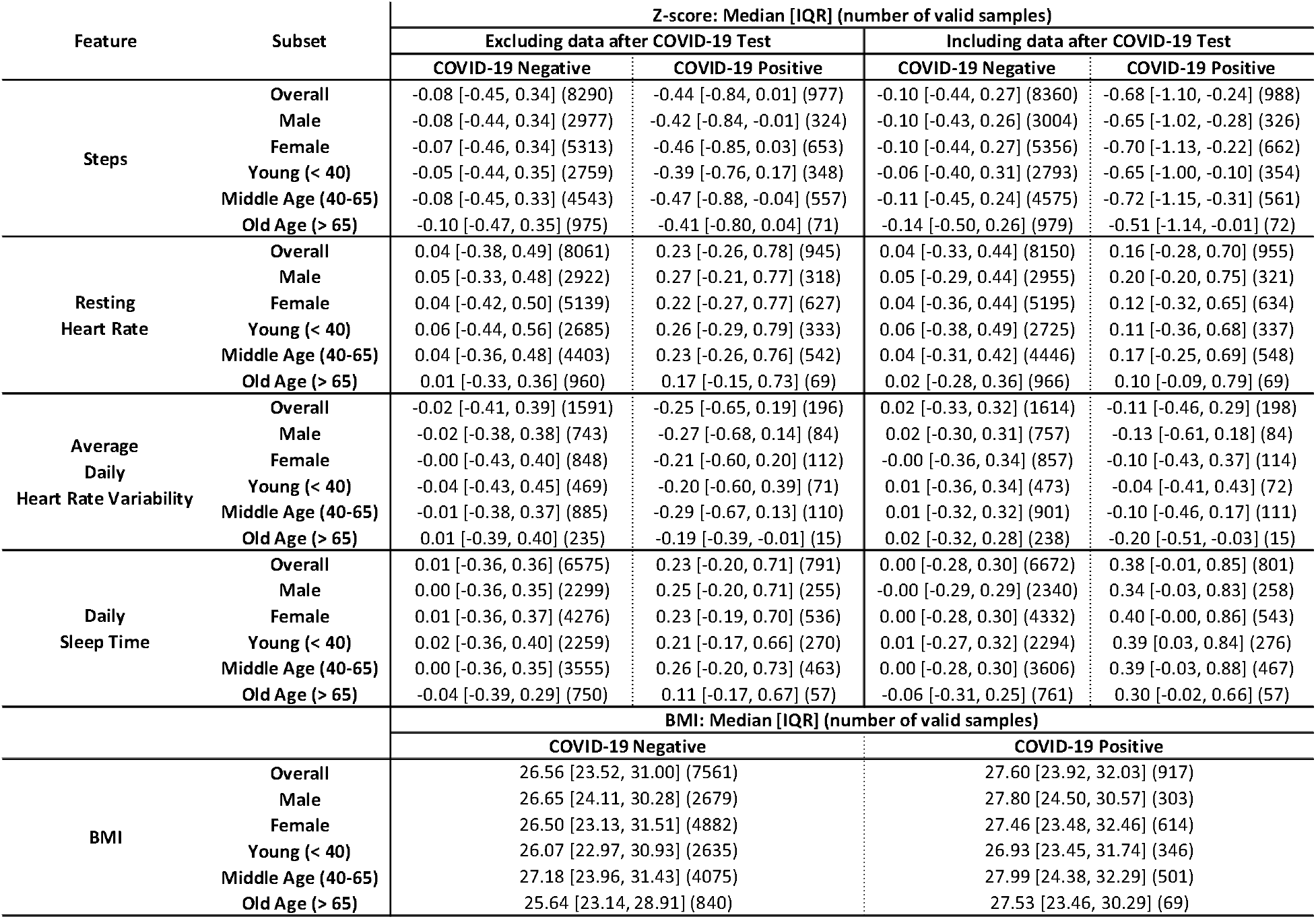
Normalized deviation from baseline values for a selected number of representative features. Values for positive and negative COVID-19 individuals, including or excluding the period after the test date, are reported. The results are stratified among gender and age groups. Median, interquartile range (IQR) and number of COVID-19 cases analyzed are reported.

**Figure 1 –.**
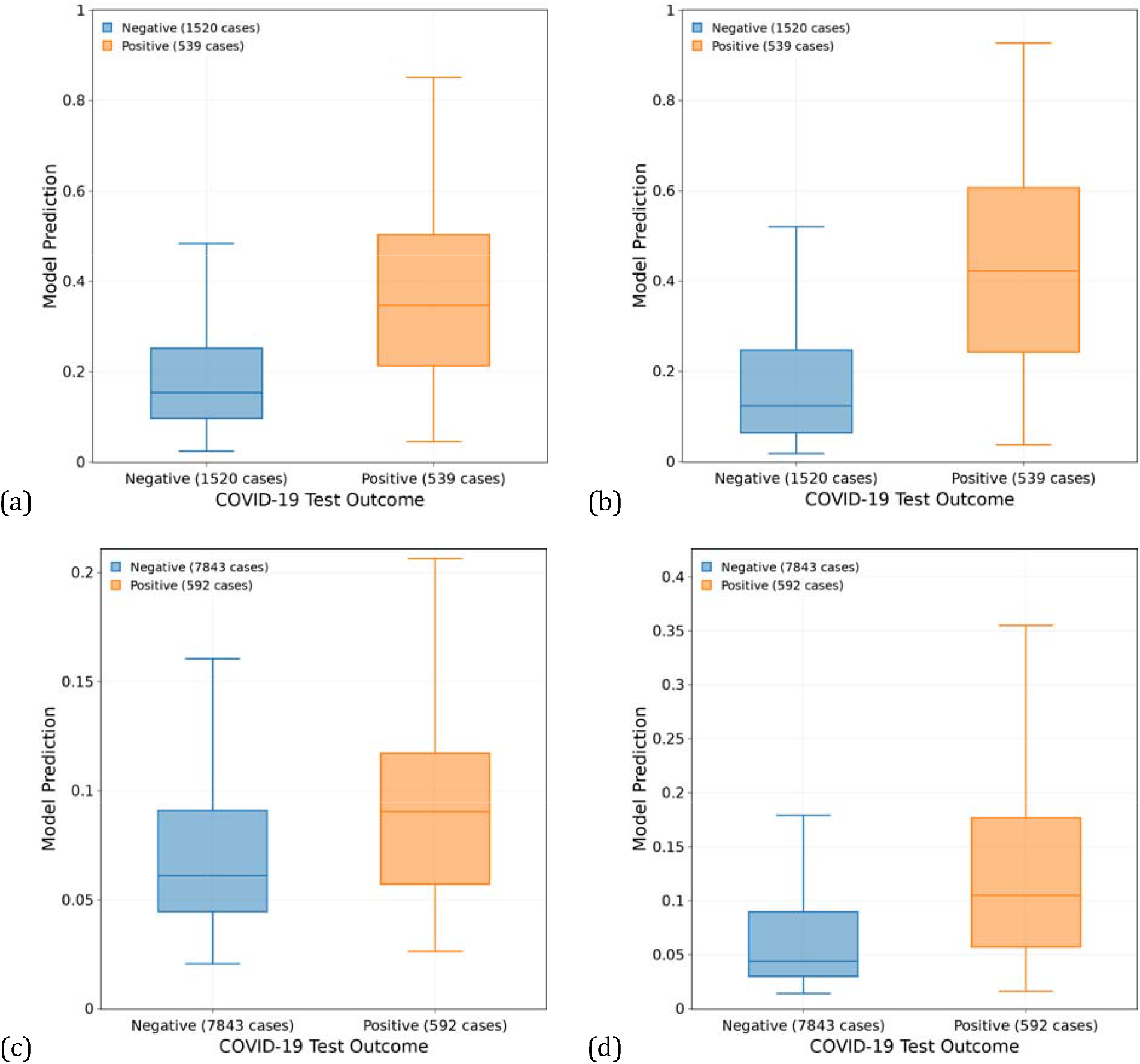
Output of the prediction models for symptomatic cases, excluding (a) and including the data after the test date (b), and for no-symptom-reported cases, excluding (c) and including the data after the test date (d). The boxes represent the IQR, and the horizontal lines are the median values. Th number of cases considered for the analysis are reported in the legend.

For the symptomatic cohort, we observed a significant difference in the model’s output between participants who tested positive or negative, showing that the two groups can be effectively separated, (Figure 1.b) even if we consider only the days preceding the test date (Figure 1.a) thus excluding any behavioral bias potentially caused by taking the test and awaiting results or knowledge of the test outcome. We showed also the predictions for the no-symptom-reported cohort, considering the data before the test date or all the available data, respectively. (Figure 1.c and 1.d) As expected, while a significant difference between the individuals testing positive or negative could still be observed, it is harder to clearly separate the two groups.

Symptomatic cases exhibited an area under the receiver operating characteristic (ROC) curve (AUC) of 0.83 [0.81-0.85], while when considering only data before the test date the performance slightly decreased, with AUC=0.78 [0.75-0.80]. For the no-symptom-reported cohort, we observed an AUC of 0.74 [0.72-0.76], or AUC=0.66 [0.64-0.68] when considering only data before the test date. (Figure 2)

**Figure 2 -.**
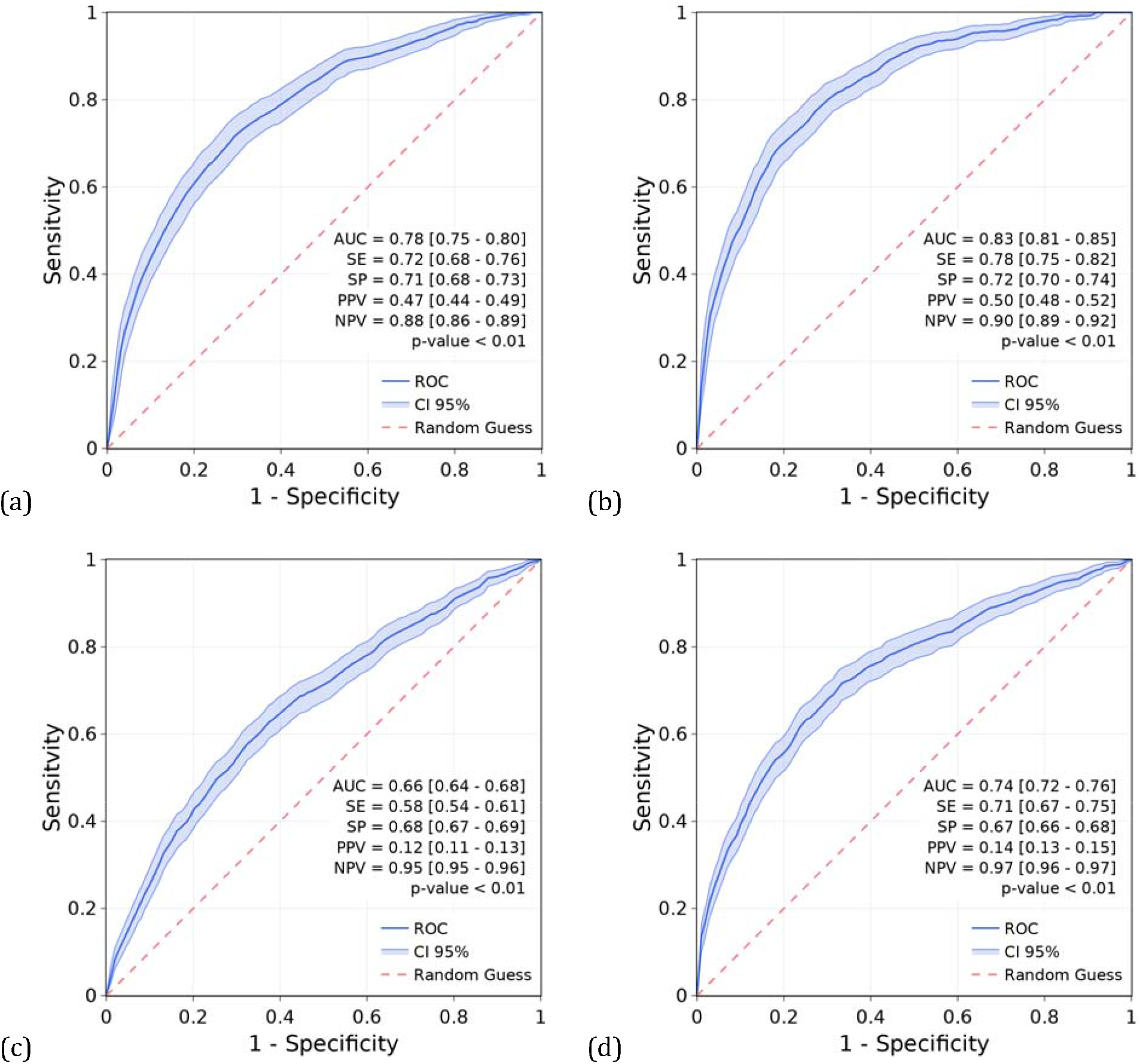
Receiver operating characteristic curves (ROCs) for the discrimination between COVID-19 positive and COVID-19 negative. Performance for symptomatic cases, excluding (a) and including th data after the test date (b), and for no-symptom-reported cases, excluding (c) and including the data after the test date (d), are reported. The model is a gradient boosting prediction model based on decision trees. Median values and 95% confidence intervals (CIs) for sensitivity (SE), specificity (SP), positive predictive value (PPV) and negative predictive value (NPV) are reported, considering the point on the ROC with the highest average value of sensitivity and specificity. Error bars represent 95% CIs. P-values of the one-sided Mann-Whitney U test are reported.

### Importance of each feature

For the symptomatic cohort, self-reported symptoms were of crucial importance for the most accurate diagnosis of the disease. Considering only data before the test, self-reported symptoms accounted for 60% of the relative contribution to the predictive model, (Figure 3.a) while considering all peri-test data, the importance of the self-reported symptoms decreased to a relative contribution of 46%. (Figure 3.b)

**Figure 3 –.**
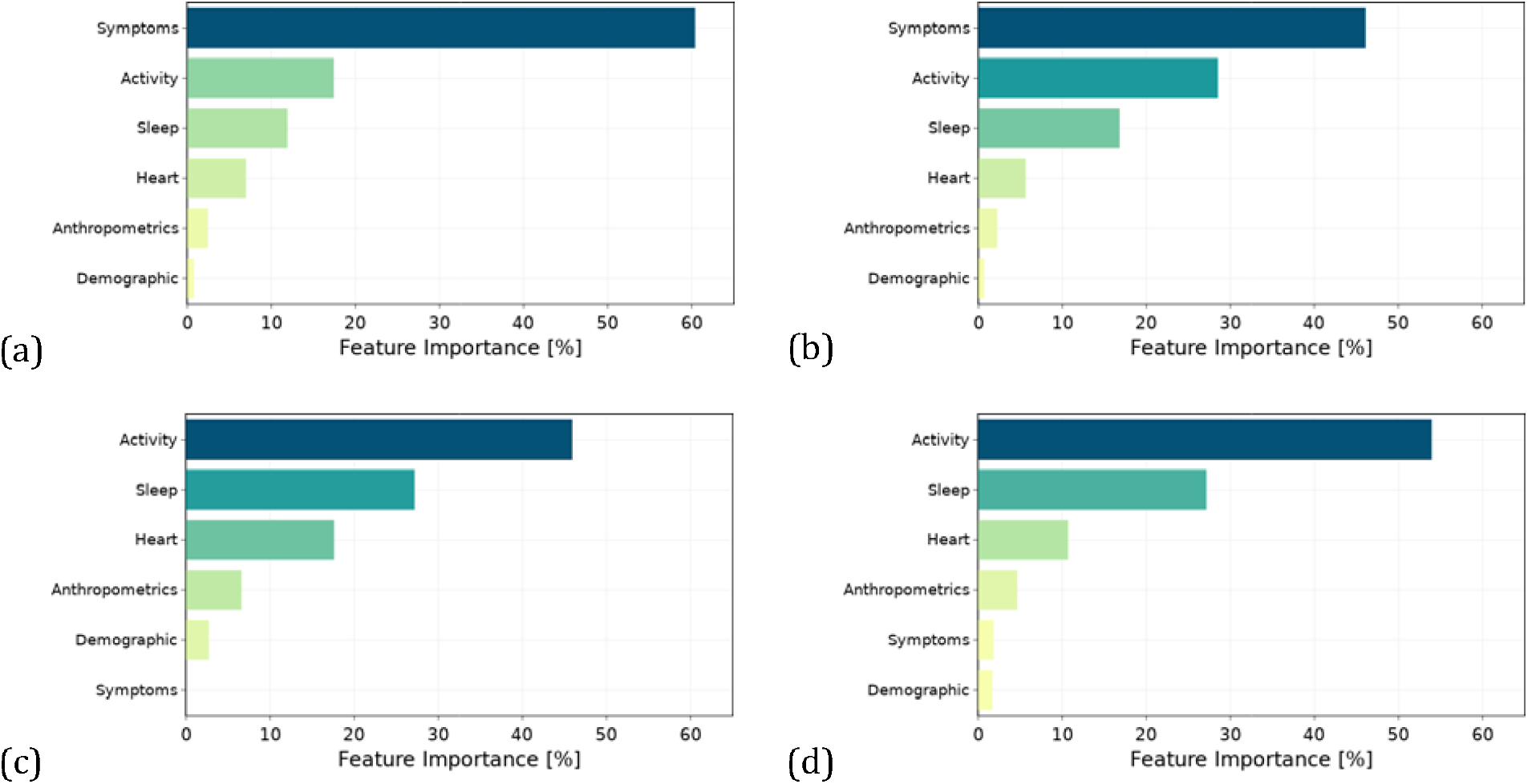
Overall feature importance based on the average prediction changes when the feature value is perturbed. Values are normalized as percentages. Features have been aggregated into macro categories. Results for symptomatic cases, excluding (a) and including data after test date (b), and fo no-symptom-reported cases, excluding (c) and including data after test date (d), are reported.

For both the symptomatic and no-symptom-reported cohorts, we observed a consistent change in the importance of the activity sensor features, if we consider only data before the test. For the no-symptom-reported cohort, (Figure 3.c and 3.d) the importance of the activity sensor features increased from 46% to 54% when all peri-test data were considered – potentially as a consequence of precautionary measures imposed after testing and awaiting results or receiving a positive test outcome. Sleep sensor features importance did not change significantly when post-test data were included for either the cohort reporting symptoms, or those not reporting symptoms, potentially because sleep was less affected by the knowledge of a test result. Sensor features in the heart rate category had a small relative contribution (6%) for the symptomatic cohort, (Figure 3.a) while their contribution increased (18%) in the no-symptom-reported cohort, (Figure 3.c) acquiring more importance in the absence of information about symptoms and when only pre-test data was considered. Anthropometrics, such as height or weight, provided only a small relative contribution, while the contribution of demographic features, such as age or gender, was negligible.

Finally, we provided more details about specific symptoms, and how each of them, on average, affects the model’s prediction. (Figure 4) We identified highly discriminative symptoms (cough and decrease in taste and smell, with ≥ 10% relative contribution), medium discriminative symptoms (congestion or runny nose, fever, chills or sweating and congestion or runny nose with <10% and ≥ 5% relative contribution) and low discriminative features (e.g. body aches, headache, fatigue, with < 5% relative contribution).

**Figure 4 -.**
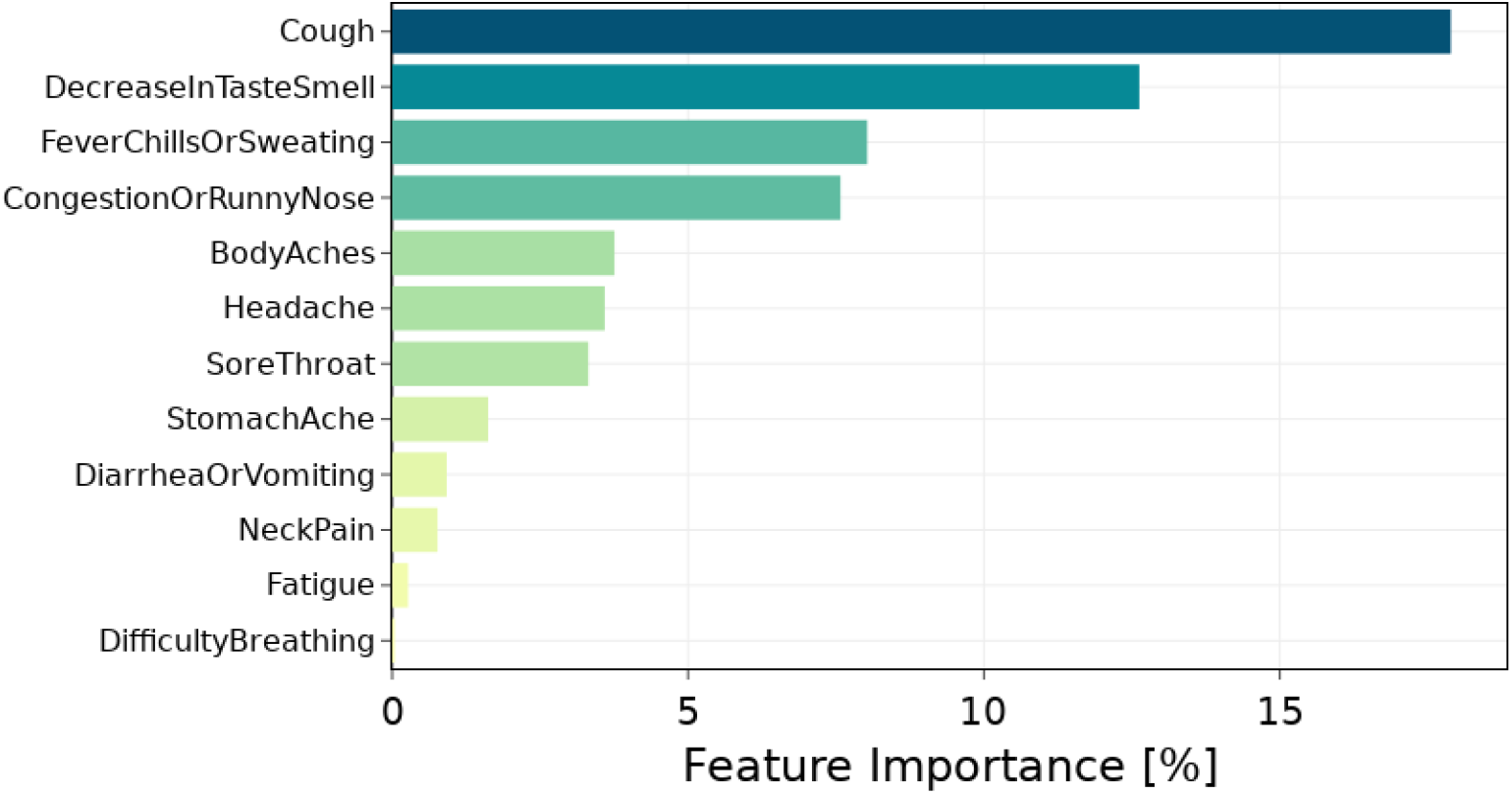
Feature importance associated to specific symptoms. Only symptoms reported before the test date have been considered. Values are normalized as percentages. The results refer to symptomatic cases only.

## DISCUSSION

Our machine learning model based on decision trees can discriminate between individuals who tested positive or negative for COVID-19 based on multiple data types collected by wearable devices, demographic information and self-reported symptoms when available. The adaptability of the algorithm to the available data allows us to also study the performance of the algorithm for individuals in the absence of self-reported symptoms, who may account for almost half of COVID-19 positive individuals.^4^ In order to estimate the effects of the behavioral changes due to the act of testing and/or receiving a positive COVID-19 test, we performed a temporal analysis dividing the data collected before and after the date of COVID-19 testing. The model has been shown to perform well for the identification of COVID-19 infection when incorporating data from symptomatic individuals that includes the five days following the date of testing, with an AUC of 0.83 (IQR: 0.81-0.85). By considering only data preceding the test date, we achieved an AUC of 0.78 (IQR: 0.75-0.80) for people who reported symptoms. When available, self-reported symptoms remain the predominant feature category considered by the model in all our test scenarios, demonstrating the importance of an engaging system that allows participants to easily report this information at any time. Among participants with symptoms, we identified cough and decrease in taste and smell as the most highly discriminative symptoms for a COVID-19 infection, followed in order of importance by fever, chills or sweating, and congestion or runny nose.

Using the same model, we also investigated individuals who did not report any symptoms. Despite the lack of self-reported information about the sickness, the model achieved an AUC of 0.74 (IQR: 0.72-0.76) when considering the period following the test, and an AUC of 0.66 (IQR: 0.64-0.68) excluding post-test data. Looking at the importance of the features used by the algorithm, we noticed that the importance of sensor-based Activity substantially increases when considering also post-test data, likely reflecting a potential behavioral change for the participants due to imposed precautionary measures.^13^ On the other hand, the importance of the heart rate features, which are less likely to be affected by short term behavioral changes, is higher when the model consider only data before the COVID-19 test. Moreover, since heart rate elevation might serve as an indicator of inflammatory conditions,^14^ its relative importance increases significantly in the absence of self-reported symptoms.

These results build on our prior retrospective work on resting heart rate^5^ and sleep,^6^ which when aggregated at the population level, have been shown to significantly improve real-time predictions for influenza-like illness.^15^ In an early study, using the initial data from DETECT, we demonstrated the potential of using self-reported symptoms and wearable data for the discrimination of positive and negative cases of COVID-19,^7^ which has been validated by several subsequent independent studies evaluating detection of COVID-19 from wearable devices.^16^ The availability of high frequency intra-day data has shown promise to identify pre-symptomatic infection^8^, even if additional studies with a larger number of individuals are needed to prove this point. Several studies focused on the specific data provided by a single sensor brand, showing that an increase in respiratory rate^17^ and heart rate,^9^ or a decrease in heart rate variability,^12^ are significant during an illness, and that the changes in these physiological signals are more severe for COVID-19 positive cases relative to those affected by other influenza-like-illnesses.^11^ We believe a strength of this research program is that anyone, with any wearable sensor, can participate. As wearable sensors continue to evolve and increase in number, predictive algorithms not dependent on a specific device or data type are needed to optimize the value of continuous, individual data. The algorithm proposed in this work is designed to ingest all available data, exploiting the information provided by the most advanced sensors, while detecting the presence of a COVID-19 infection for everybody owning any type of wearable sensor. The algorithm recognized the importance of self-reported symptoms in the prediction accuracy, but it is also designed to work in the absence of them, thus extending its applicability to the asymptomatic, pre-symptomatic or just a less engaged population who may not want to bother with reporting symptoms.

The analysis of individuals without self-reported symptoms extends the use of the algorithms for a fully passive monitoring of the pandemic and provides the possibility of applying the algorithm in other settings that collect wearable sensor data but are not equipped to collect and analyze self-reported symptoms. (Supplementary Material) Among them, the largest is Corona-Dataspende, a project developed by the Robert Koch Institute to collect sensor data from more than 600,000 individuals, monitoring the course of the pandemic in Germany.^18^

The negligible importance given by our algorithm to the demographic features may be explained by observing that the physiological features we consider are changes with respect to an individual baseline. While an individual’s baseline differs based on their demographic features, the changes with respect to the baseline that we use in our algorithm are not much affected by the demographic characteristics of the individual.

While the use of machine learning in the detection and prognostication for COVID-19 based on chest radiographs and CT scans have been questioned in a systematic review that discussed how none of the current studies are of potential clinical use due to biases or methodological flaws,^19^ the use of machine learning to enable a continuous and passive COVID-19 early detection is both very promising -for the potential to be scaled up effectively to a large fraction of the population -and repeatable -since we used a strictly separated test set for each of the cross-validation folds. Furthermore, the prediction algorithms developed as part of the DETECT system could be adapted to study the long term health problems due to COVID-19,^20-24^ or the effects of COVID-19 vaccine on vital signs and individual behavior.^25-27^ For future infectious pathogen epidemics and pandemics, the new machine learning algorithms developed from the DETECT data can be adapted and re-used for early detection of various types of infections, towards the development of a new system to monitor the spread of future viral illness and prevent future outbreaks or pandemics.

### Limitations

In DETECT, all data is participant reported with no validation of the accuracy of self-reported symptoms, test dates or results. While we were able to collect continuous data, the amount of sensor data collected, or the accuracy of self-reported symptoms, depends entirely on the willingness of the participants to wear the sensor and accurately report how they feel. Despite the fact that the information collected may not be as accurate as in a controlled laboratory setting, previous work has demonstrated the value of participant-reported outcomes.^28-30^ In the data analysis, among the people who reported the COVID-19 test outcome (active participants), we separated participants who reported at least one symptom from those who did not report any symptoms. The app indeed did not have an explicit way to report the absence of symptoms, so potentially some symptomatic individual may have not reported their symptoms.

Furthermore, this study is based on the aggregation of continuously monitored data into a finite number of daily features. A recent study has provided new insights about the analysis of intra-day changes for monitoring physiological variations,^31^ that may be used in future studies. Changes in more advanced metrics, like respiratory rate,^17^ peripheral temperature^10^ or HRV,^12^ may also prove to add to the prediction of a COVID-19 infection, even if they have been marginally considered in our work since only a small fraction of participants were providing this type of data.

While previous studies have shown the importance of remote monitoring of individuals, extending health research beyond the limits of brick and mortar health systems,^32,33^ additional disparities are introduced when the study relies on wearable sensors, due to reduced accuracy for certain skin tones^34^ and unequal access to this digital technology.^35^ The decreasing cost of wearable sensors (some now less than $35) and the inner adaptability of our detection algorithm to any sensor and any given level of engagement of the participant with the in-app system will hopefully help in decreasing the barriers for underserved and underrepresented populations.

## METHODS

### Study Population

Individuals living in the United States and at least 18 years old are eligible to participate in the DETECT study. After downloading the iOS or Android research app, MyDataHelps, and consenting into the study, participants are asked to share their personal device data (including historical data collected prior to enrollment) from any wearable device connected through direct API (for Fitbit devices), or via Apple HealthKit or GoogleFit data aggregators. A participant is invited to report symptoms, diagnostic test results, vaccine status, and connect their electronic health records, but they can opt to share as much or as little data as they would like. The recruitment of participants happens via the study website (www.detectstudy.org), several media reports, or outreach from our partners at Walgreens, CVS/Aetna, Fitbit and others.

### Ethical Considerations

All individuals participating in the study provided informed consent electronically. The protocol for this study was reviewed and approved by the Scripps Office for the Protection of Research Subjects (IRB 20–7531).

### Data collection, aggregation, and group definition

All the participants with at least one self-reported result for a COVID-19 swab test during the entire data collection period have been considered in this study. Based on the reported data, an individual is considered Negative if the test resulted negative and no other positive tests have been reported in the period from 60 days before to 60 days after the test date. A minimum distance of 60 days is guaranteed between tests from the same individual considered in the analysis. This ensures that, if multiple tests are reported in the same period, only the first one is considered in our analysis, and the ones reported in the following 60 days will be ignored.

For each participant, we collect the data preceding and following the test date from all the connected devices, including, among others, detailed sleep intervals, number of steps and daily resting heart rate values. All the considered metrics are reported and detailed. (Table 1) If multiple values per day are available for the same data type, a specific pre-processing has been applied to obtain a single representative daily value. Data has been collected from all the devices synchronized with the Fitbit or HealthKit application available on the smartphone. If data of the same type is available from multiple devices, only the most used device in the monitored period is considered.

Along with device data, we also analyze the reported surveys looking for self-reported symptoms. We considered all the symptoms reported from 15 days before to the day of test, further dividing the participants into two groups: Symptomatic cohort, if we observe at least one reported symptom before the day of test, and non-symptom reporting cohort, if no symptom has been reported during this period. The frequency of each reported symptom for positive and negative cases are also reported. (Figure 5)

**Figure 5 –.**
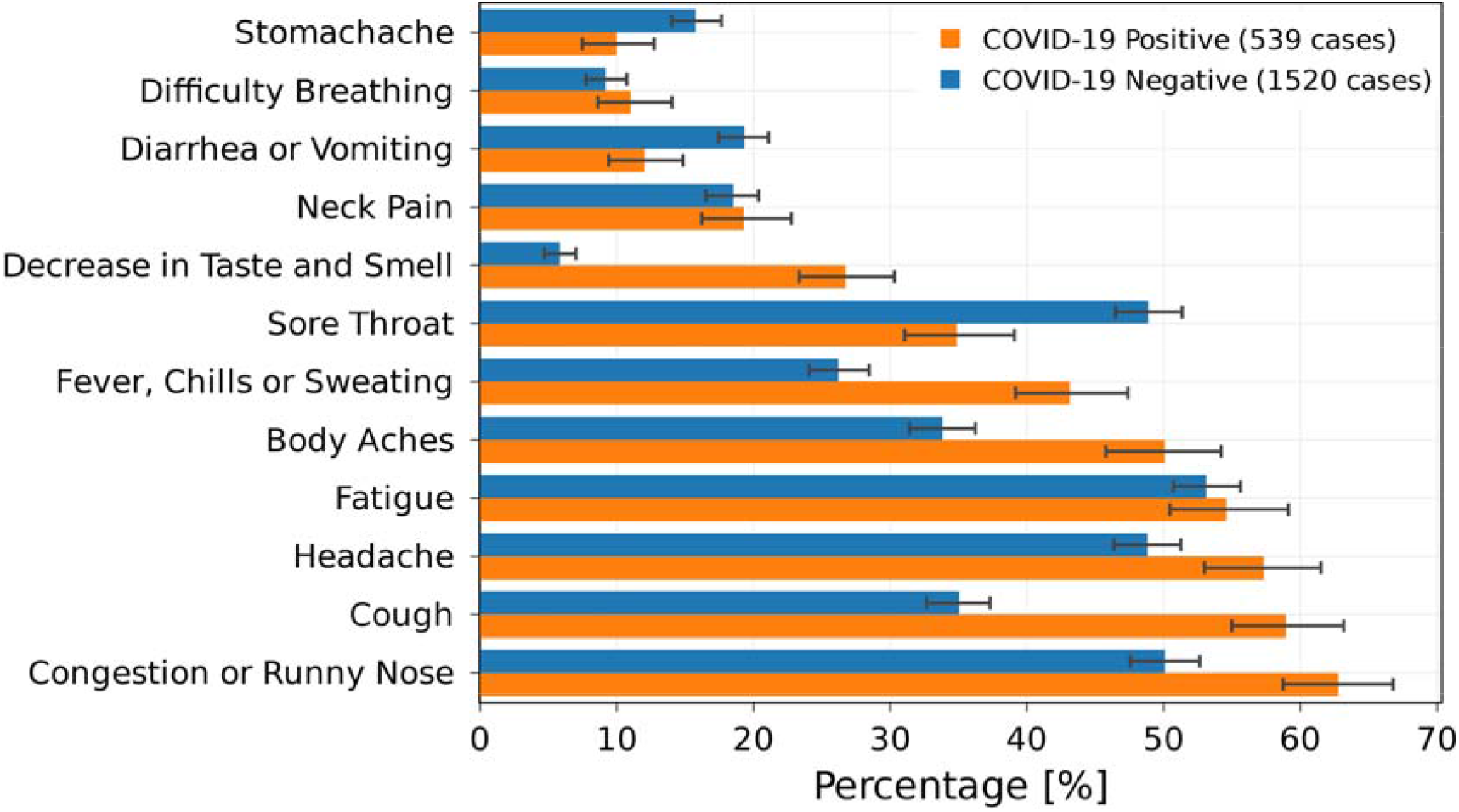
Percentage of reported symptoms for participants who reported at least one symptom from 15 days before to 5 days after the test date. The frequencies of the indicated symptoms are shown for positive and negative cases. The error bars represent 95% percent confidence intervals.

### Baseline evaluation

Behavioral and physiological data acquired from wearable devices are highly subjective. The intrinsic inter-individual variability of physiological metrics, the different habits of the users, and the multiple purposes of the wearable devices requires a careful definition of the subjective baseline value for each of the considered metrics. Thus, the daily baseline is calculated using an exponentially weighted moving average:

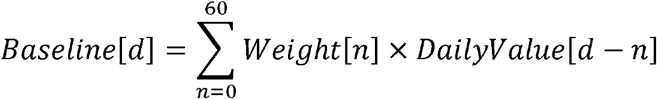

where *d* is the current day and *n* represent the number of days before *d*, with a maximum of 60 days before the current date, while *DailyValue* can be any of the daily data measures among the ones considered.

The oscillation of the measures during the baseline period is also subjective to change over time. To measure the daily baseline variability, we evaluate the weighted standard deviation using the same weights of the baseline

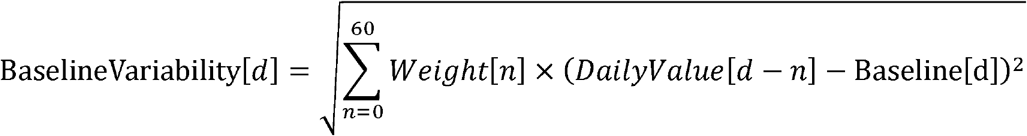

The weights (Figure 6) decrease exponentially as *e*^-*an*^ with *α* =0.05. We exclude the first (*n* < 7) days from the computation to avoid recent changes to affect the baseline value.

**Figure 6 –.**
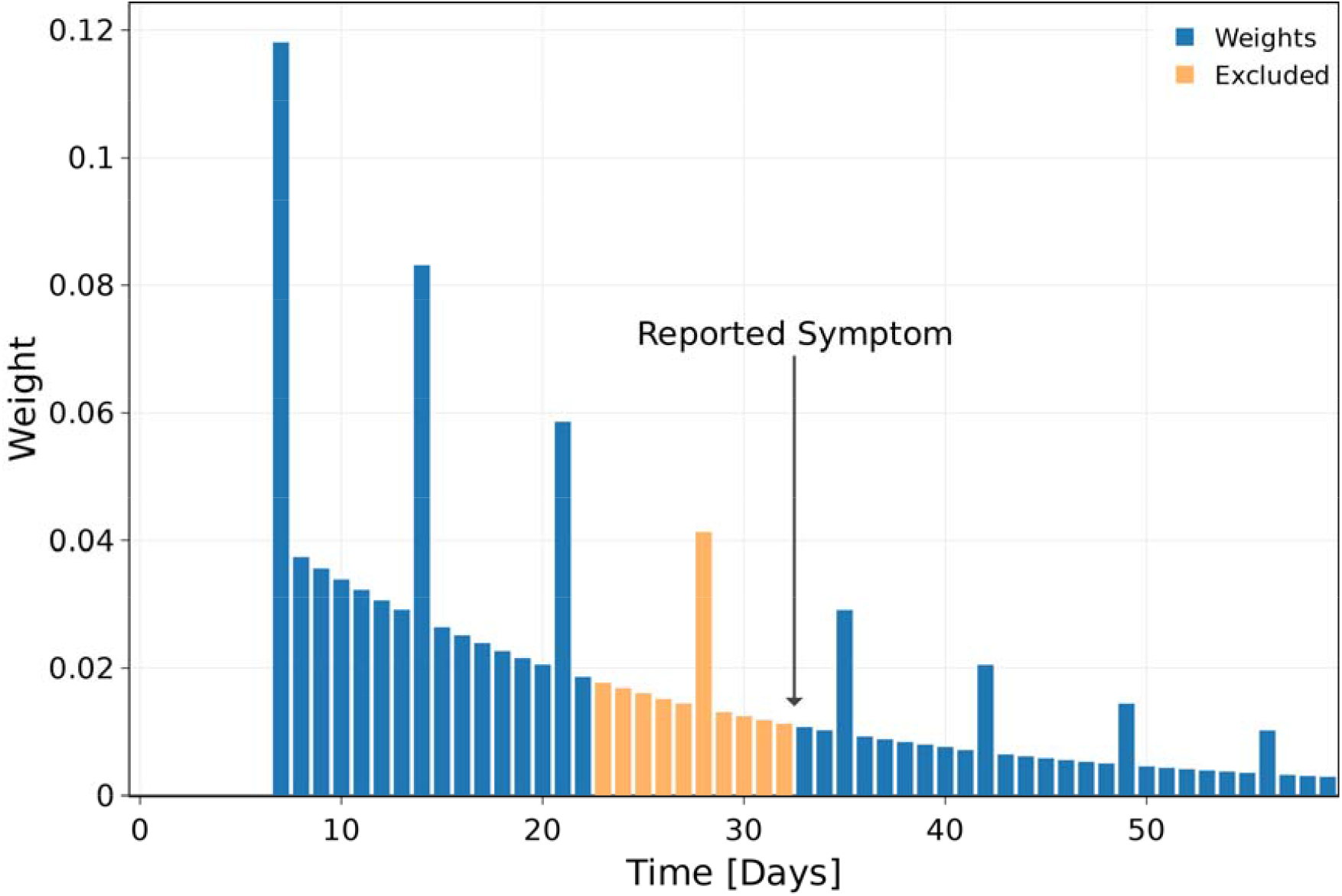
Exponentially decreasing weights for the evaluation of the baseline data, with weekly patterns. The abscissa represents the temporal distance preceding the analyzed day considered for th baseline evaluation. The first 6 days have been excluded to avoid recent changes to affect the baseline. Additionally, if a symptom has been reported in this time frame, we set the weights to zero from the day of symptom to the next 10 days.

Many behavioral habits present strong weekly patterns, such as an increased sleep duration during the weekend, or weekly physical activities. To take into account these behaviors in the baseline evaluation, and to reduce the chances of false positives, we consider weekly patterns by increasing three times all the weights corresponding to the same day of the week.

During the course of a temporary disease, physiological measures may be different. In order not to affect the baseline value, which should only depend on the normal behavior, we exclude the 10 days following any reported symptoms from the baseline evaluation.

Finally, the weights are normalized to sum to 1.

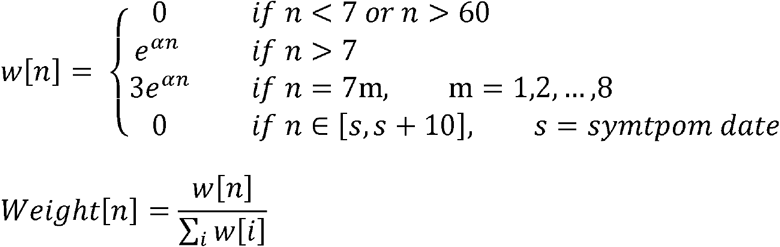

The deviation from the baseline values is then evaluated as:

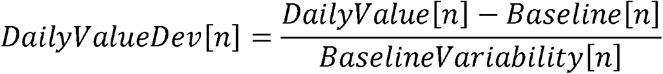

This value represents how far the specific metric is from the expected normal value, day by day. Values are considered to be valid only if at least 50% of corresponding data are available during the baseline period.

### Feature extraction

We propose two analyses, one considering all available data (5 days before and 5 days after the test date), and one considering only the period before the test date (5 days before test), in order to further analyze the impact of the test outcome on the individual behavior and the natural course of the disease.

We consider four different macro categories of feature. (Table 1)

– *Sensor features*: all the features acquired or derived from the device measurements belong to this group. In this study, we consider the minimum, average, and maximum deviation values from the baseline in the days considered. This category is further divided into 3 sub-categories, including activity, heart and sleep related features.
– *Symptom features*: a separate binary feature is considered for each of the reported symptoms. If the corresponding symptom has been reported in the considered period its value is set to 1, otherwise 0.
– *Anthropometrics*: if available from the monitored devices, several anthropometric features are also considered like body weight, height, body mass index, fat percentage, and basal metabolic rate.
– *Demographic features*: this category includes age and gender self-reported by the participants.

Using the aforementioned features, we developed a gradient boosting prediction model based on decision trees^36^. The model has been trained and tested in four different conditions, using data from the symptomatic or no-symptom-reported cohort, and preceding the reported test date or considering all available data around the test date. Normalized deviation (Z-score) from a subjective and dynamic baseline value was evaluated daily for each metric and each individual. A weighted average based on past data was defined as the baseline estimation, whose weights are reported. (Figure 6)

The entire dataset has been randomly divided into 5 separate non-overlapping test sets. For each test set, a model is trained using all the remaining data, ensuring an equal percentage of positive cases between train and test sets. For each model, we also ensure that the test set remains strictly separate from the training, so training data are not involved in the test.

To analyze the intrinsic variability of the model due to data availability, we estimate 95% confidence intervals for the presented results. Bootstrap method has been utilized for this purpose, with 10,000 independent random iterations from the test outcomes.

To have a better understanding of the effect of COVID-19 on physiological and behavioral aspects, we consider symptomatic and no-symptom-reported cases separately. Additionally, different models have been analyzed considering sensor features evaluated including and excluding sensor data after the reported test date. Comparative results are presented in terms of AUC of the ROCs. Sensitivity (SE), specificity (SP), positive predictive value (PPV) and negative predictive value (NPV), associated to an optimal operating point, are also reported. The optimal operating point is defined as the point with the highest average value between SE and SP.

The interpretable nature of the decision tree model allows for the evaluation of feature importance estimates^37,38^. To this end, we evaluate, for each feature, the average prediction changes when the feature value is perturbed. The higher the change to the prediction value, the higher the contribution given by the corresponding feature to the model’s outcome. To have a more comprehensive overview of the feature importance, we further aggregated the importance associated to features in the same category.

## Data Availability

This is an ongoing study. We plan to make the de-identified data available after approval of a proposal by a responsible authority at Scripps and with a data access agreement, pledging to not re-identify individuals or share the data with a third party.

## ACKNOWLEDGEMENTS

This work was funded by grant number UL1TR002550 from the National Center for Advancing Translational Sciences (NCATS) at the National Institutes of Health (NIH) (E.J.T., S.R.S., G. Q.) and by grant number OIA-2040727 (Convergence Accelerator) at the National Science Foundation (NSF) (M.G., G.Q.).

## AUTHORS CONTRIBUTIONS

M.G., and G.Q. made substantial contributions to the study conception and design. M.G., J.M.R., K.B.-M., E.R., S.R.S, and G.Q. made substantial contributions to the acquisition of data. M.G., and G.Q. conducted statistical analysis. M.G., S.R.S., and G.Q. made substantial contributions to the interpretation of data. M.G., and G.Q. drafted the first version of the manuscript. M.G., J.M.R., K.B.-M., E.R., V. K., E.J.T., S.R.S., and G.Q. contributed to critical revisions and approved the final version of the manuscript. M.G., and G.Q. take responsibility for the integrity of the work.

## SUPPLEMENTARY MATERIAL

### Analysis in the absence of self-reported symptoms

For an improved and effective tracking of the pandemic, many research institutions are collecting sensor data from individuals, while it is not always possible to provide surveys and collect active feedback from participants. Information actively added by participants may be crucial especially for potentially infected individuals, indeed a fully passive data collection system may be adopted by a broader audience and have a more capillary diffusion.

Our model can be leveraged to support also these studies based uniquely on passive data collection. To this end, we performed an additional analysis without the inclusion of self-reported symptoms as source of knowledge for the model. The results of the analysis without considering self-reported symptoms are shown in terms of AUC of the ROC. (Figure S.1)

**Figure S.1.**
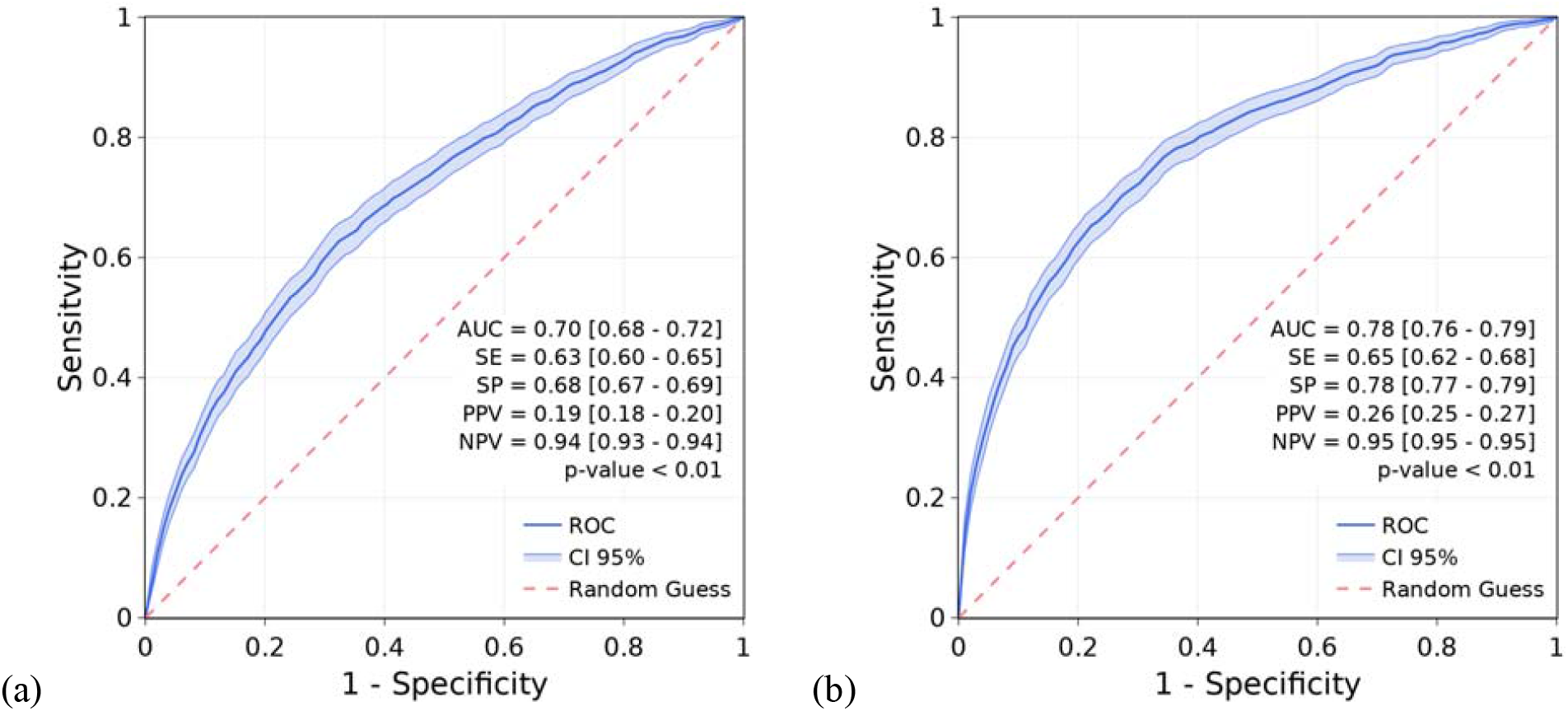
Receiver operating characteristic curves (ROCs) for the discrimination between COVID-19 positive and COVID-19 negative when the self-reported symptom data is not used in the prediction. Performance obtained by excluding (a) and including data after the test date (b) are reported. The model is a gradient boosting prediction model based on decision trees. Median values and 95% confidence intervals (CIs) for sensitivity (SE), specificity (SP), positive predictive value (PPV) and negative predictive value (NPV) are reported, considering the point on the ROC with the highest average value of sensitivity and specificity. Error bars represent 95% CIs. p-values of the one-sided Mann-Whitney U test are reported.

